# Measurement of subjective and objective indices after progressive resistance training compared with aerobic training in Patients with Haemophilia: a study protocol

**DOI:** 10.1101/2022.01.27.22269981

**Authors:** Felicianus Anthony Pereira, Nabila Najam Soomro, Farheen Sulaiman

**Affiliations:** National Institute of Physical Therapy & Rehabilitation Sciences; Sindh Institute of Physical Medicine and Rehabilitation

**Keywords:** Hemophilia, resistance training, aerobic exercise, Pakistan

## Abstract

**Background:** Bleeding episodes in mild haemophilia may occur after major injuries or surgical procedures with some people not experiencing bleeding episodes. People suffering from moderate haemophilia bleed once a month, however they rarely experience spontaneous bleeding. Those suffering from severe haemophilia bleed quite often into muscles or joints, and episodes can occur once to twice a week. Bleeding usually occurs spontaneously.

**Objective:** To investigate the effects of progressive resistance training on quality of life, muscular strength and joint score in patients with haemophilia.

**Methodology:** Sixty patients will be enrolled in the study. Thirty patients will be allocated to control group, and thirty to intervention group. Controls will be administered active muscle stretching and aerobic exercises. Intervention group will be given active muscle stretching, and resistance training. Patients will be randomly allocated to each group. Anthropometric data will be measured pre-test to establish a baseline. Study variables include muscular strength, and quality of life. All tests will be measured pre-test and post-test to compare effects of treatment.

**Results:** Participant recruitment commenced in June 2021. The post intervention phase will be completed by August 2020. Data analysis will commence after this. A write-up for publication is expected to be completed after the follow-up phase is finalized in August 2021.

**Conclusions:** If resistance training is found to be effective in improving quality of life and muscular strength in participants, it could reduce the frequency of factor therapy given prophylactically, or even as acute treatment, thus directing it towards more severe cases. It will also provide financial relief to organizations supporting the treatment of the hemophilic population.

## 1. INTRODUCTION

Haemophilia is an X-linked congenital bleeding disease which occurs due to lack coagulation factors VIII or factor IX. This deficit is an effect of mutations in clotting factor genes (FVIII in type A, and FIX in type B). Occurrence of haemophilia is approximately 1 in 10 000 births.^1^ Haemophilia A is found more commonly in the population. Regardless of the type of haemophilia, the outcome is the same; people affected bleed longer than normal. In the healthy population, normal international units (IU) are 0.50-1.50 per milliliter of whole blood. People with mild haemophilia have levels of 0.05-0.4 IU, and people with moderate haemophilia have levels of 0.01-0.05 IU. Severe is below 0.01 IU.^2^ People with mild haemophilia might experience bleeding after surgical procedures or major injury, while others may not experience bleeding episodes. People with moderate haemophilia bleed around once a month and rarely experience spontaneous bleeding. Those suffering from severe haemophilia bleed quite often into muscles or joints, and episodes might occur once or twice a week. Bleeding usually occurs spontaneously.^2^ In the annual global survey conducted across 116 countries by World Federation of Hemophilia (WFH) in 2017, a total of 196,706 people have been identified with haemophilia. In Pakistan, 1743 people were diagnosed with haemophilia, 250 had Von Willebrand’s disease, and 121 had other associated bleeding disorders.^3^ Psychosocial factors play a key role in quality of life (QoL) of patients with haemophilia (PwH).^4,5^ As severity of hemophilia increases, the QoL in PwH decreases when compared with healthy population.^6^ Self-esteem of males who have severe bleeding disorders is also considerably worse compared to that of their healthy peers, whereas females show significant differences. ^7^ Treatment for haemophilia is via factor replacement therapy, which may be given prophylactically, or as acute treatment.^8^ Hemophilic arthropathy can be extremely painful, with knees, elbows and ankle most commonly being affected. Recurring bleeding in the joint can cause synovitis that can develop into arthropathy, which may later disrupt activities of daily living. The joint most commonly affected is the knee, with a simple act such as ambulating on level surfaces generating substantial force and stresses on the knee joint. Due to muscle weakness, the knee joint is further susceptible to weight bearing stresses, which results in a loop of persistent joint bleeds and rising synovitis which causes further muscle atrophy, eventually leading to severe arthropathy. ^9^ Traditionally PwH were often advised against participating in sports due to possibility of injury, however, studies have tried to demonstrate the positive effects of exercise programs for PwH.

## Methods

The study design is a single blind, randomized controlled trial, and study setting is single center study. Ethical approval has been obtained from institutional review board (Ref no. IRB-1936/DUHS/Approval/2021/), and has been registered as a clinical trial in clinicaltrials.gov on 19^th^ May, 2021 (identifier: NCT04892628). Sample size was calculated using OpenEpi version 3.0 calculator. In the section of overall HJHS score of resistance training group ^(14)^, the detectable difference and clinically important difference ranged from 7.25±1.28 and 4.83±1.19 points. A power of 80% and confidence interval of 95% was set and a sample size of 5 per group was obtained. However, the sample size will be increased to 30 participants per group.

### Inclusion Criteria

The study will include participants:

- clinically diagnosed with hemophilia A or B (mild to moderate hemophilia (mild hemophilia - 0.05-0.4 IU, moderate hemophilia - 0.01-0.05 IU)
- willing to undergo pre- and post-program objective assessment of manual muscle testing and Hemophilia Joint Health Score, as well as subjective assessment of and Rating of Perceived Exertion.
- willing to train two times a week
- males and females between 18 to 45 years of age.
- those who can ambulate without assistance

### Exclusion Criteria

- unable to attend exercise sessions for the complete duration of the study
- those who have had surgery performed in the past six weeks, or are scheduled for surgery in duration of training
- involvement in any other training, or rehabilitation, during study
- change in medicine within the study
- history of major bleeding episodes that could pose a risk
- History of FVIII or FIX inhibitor

#### Data collection

Data will be collected from a Hemophilia Welfare center located in Karachi, Pakistan. A hematologist, orthopedic and general physician, as well as paramedical staff are present on-site, to guarantee safety of patients. Sixty participants will be included in the study. Written informed consent will be obtained from participants, with forms being provided in both English and in Urdu. Once consent has been obtained, participants will be enrolled in the study. Participants will be divided equally in two groups: a control group and an intervention group. Allocation to both these groups will be done by simple random sampling, using a computer software (randomizer.org), with participants unaware regarding group allocation. Participant data is available via the registry maintained by the welfare center, providing a sample frame from which participants can be selected.

#### Intervention

Participant’s height, weight and age will be recorded for pre-test anthropometric data. Height will be measured using a stadiometer, and weight will be measured with an electronic scale. Hemophilia Joint Health Score v2.1 and Manual Muscle Testing (Daniels and Worthingham’s) for deltoid (anterior, middle and posterior fibers), biceps, triceps, quadriceps, hamstrings, and calf muscles will be conducted pre and post-test. HEP-Test-Q will be assessed pre and post-test to note change in subjective QoL. Rating of perceived exertion (Borg rating of perceived exertion) will be assessed on every session to check progression in variables being tested. HEP-Test-Q and Rating of perceived exertion will be provided in Urdu for participants. Treatment sessions will be conducted twice a week, for eight weeks, amounting to a total of sixteen sessions. The control group will be administered a standard physical therapy intervention program which will consist of active muscle stretching and aerobic exercise. License has been obtained from the relevant authorities to use these questionnaires in this study.

Exercises will include:

- Flexion, extension and abduction at the shoulder joint for anterior, middle and posterior fibers of deltoid respectively
- Flexion and extension at the elbow for biceps brachii and triceps trachii
- Flexion and extension at the knee joint for quadriceps and hamstring muscles
- Plantar flexion at the ankle joint for gastrocnemius and soleus

Three sets of ten repetitions of each exercise will be done.

The intervention group will undergo a progressive resistance training program consisting of active muscle stretching and resistance training. Exercise for intervention group will consist of:

- Resisted flexion, extension and abduction at the shoulder joint for anterior, middle and posterior deltoid, using dumbbells
- Resisted flexion and extension at the elbow for biceps brachii and triceps trachii, using dumbbells
- Resisted flexion and extension of knee joint for quadriceps and hamstring muscles, using therabands
- Resisted dorsiflexion and plantar flexion of gastrocnemius and soleus, using therabands

Resistance will be increased on basis of progressive overload principle. Three sets of ten repetitions will be performed per muscle group. Initial weight of the dumbbells will be 1kg and will be increased by 0.5kg per week. Therabands used will be Yellow, Red, Green and Blue. Resistance for therabands will be upgraded every two weeks. Treatment time per group will be approximately forty minutes. Exercises will be progressed in a gradual manner to give participants time to build muscle strength, and prevent chances of injury.

#### Statistical Treatment

##### Study variables include

- Progressive resistance training, gender and age, weight are the independent variable
- Quality of life, muscle strength, rating of perceived exertion and joint score are the dependant variables

### Statistical Analysis

The software for data analysis will be IBM SPSS version 21. Mean and standard deviation will be calculated for qualitative variables. For quantitative variables, paired sample T-test will be applied.

Price of one injection of FVIII in Pakistan is around USD 80/-^10^. A patient who requires 12-16 injections every month, the cost will be roughly USD 11,500 ∼ USD 15,500/-anually.^10^ In a country where the yearly minimum wage is $2,484.00 USD, affording such treatment becomes difficult for the population.

#### Limitations

- Isokinetic dynamometer cannot be used to assess muscular strength due to cost of equipment.
- Questionnaires such as Haem-A-QoL that required purchasing fees cannot be used
- Study setting is a single center study

## REVIEW OF LITERATURE

A systematic review was conducted in 2015 by G. S. Schäfer et al in nine controlled clinical trials on pain and musculoskeletal function in PWH. The study found that exercise and physical therapy maneuvers decrease pain perception and increase range of motion and muscular strength in the hemophiliac population. It also noted that further RCTs, with greater emphasis on methodology and greater focus on parameters of exercise prescription are required.^11^ A systematic review on methods and results of psychosocial features of hemophilia displayed that studies into psychosocial aspects of the disease is still lacking. Most of the studies conducted are questionnaire based. Results across studies could not be assessed due to the variety of questionnaires used. PwH are affected by factors which may consist of life satisfaction, self-esteem, anxiety and depression. It was also noted that stressors could differ all through hemophilia life cycle.^4^ A systematic review on benefits of exercise for PwH found that correctly performed exercise along with involvement in suitable sports are helpful for PwH. Competing in sports provides many physical benefits, along with boosting emotional and societal well-being. The article also points out that participation should be in appropriate sports, as each case of hemophilia is unique.^12^

A clinical trial conducted in 2003, of physical training in subjects with hemophilia confirmed specific sports therapy consisting of proprioceptive utility with mild strength exercise increased the proprioceptive functioning along with muscular power.^13^ A randomized controlled trial conducted in 2016 by Runkel et al on programmed sports therapy, among 64 PwH found significant differences in values of triceps, biceps, latissimus dorsi, and quadriceps femoris between control and intervention groups. Outcome measures used in the study included strength measurements, joint score, co-ordination test and 12 minute walk test.^14^ In a study by S El-Shamy on the outcome of whole body vibration in children suffering from hemophilia found significant differences between intervention group and control group. (Quadriceps peak torque p<0.001, functional capacity p=0.006).^15^ Data obtained from a study of 32 patients by Greene et al showed that significant increase in muscular strength of flexor (p<0.0001) and extensor (p<0.001) aspect of the knee can be achieved by an isokinetic training program. Greater improvement was noticed in patients whose severity of arthropathy was lower, who displayed an increase of 1cm thigh girth, and who exercised more frequently.^9^

A RCT was conducted in 2017 on 20 patients to assess effectiveness of home exercise program in adult PwH. Significant difference was noted between QoL, illness behaviors, and pain perception of ankle; however no significant difference in joint status was noted. The authors concluded that perception of pain can be altered by administering a home exercise plan.^16^ A review article by Stephensen et al on orthopedic co-morbidities in elderly hemophilia population highlighted joint status, pain, muscle atrophy and strength, balance and gait, physical activity and QoL and bone mineral density. The study found that exercise can help slow down normal degeneration processes that occur with ageing. Recommended exercise frequency is two to three times a week, for at least 12 weeks in the ageing hemophilia population.^17^ A cross-sectional study was conducted among 12 PwH (11 severe, and 1 mild) to assess if upper body muscle strength would vary depending on the intervention. Results of the study showed that external resistance of muscles at moderate intensities, compared to conventional non-resisted therapeutic exercises, provide greater muscle activity than non-resisted exercises.^18^ An observational study on QoL in children with hemophilia and sporting activities was conducted by R. Cuesta-Barriuso et al. fifty three children with hemophilia and fifty one children without were assessed. Exercises were done bi-weekly over 15-weeks. Home exercises were done daily, 6 days a week, Outcome measure used to assess objective measures was the Spanish version of HJHS. No considerable discrepancy was found in perception of QoL among groups. Participation in sports increased the joint health score and QoL in children with hemophilia.^19^ A nationwide population-based study was conducted in Taiwan. Data was obtained from patients who were recorded in the national database. The study set out to assess factors influencing availing of rehabilitation. Total rehabilitation cost stood at <0.1% of total yearly medical expenses. Physical therapy accounted for 71.2% of rehabilitation therapy. The study concluded that surgical procedures related to nervous system and musculoskeletal systems, and higher usage of FVIII affected rehabilitation usage by PwH A (odds ratio 3.788; p<0.001).^20^ A randomized controlled trial conducted in 2012 on therapeutic exercises versus hydrotherapy on pain intensity in PwH both methods decreased pain, and increased range of motion. Hydrotherapy was more effective than exercise in management of pain. Significant difference was not found in range of motion after intervention with these methods.^21^

## Data Availability

All data produced in the present work are contained in the manuscript

